# Epidemiology and Antimicrobial Susceptibility Patterns of *Staphylococcus aureus* Isolates from Clinical Specimens at Meru Teaching and Referral Hospital, Kenya

**DOI:** 10.64898/2026.09.09.26362593

**Authors:** Pharis Kaumbuthu, Dorothy Kagendo, Jane Rutto

## Abstract

**Background:** *Staphylococcus aureus* is an important bacterial pathogen responsible for a wide range of community- and healthcare-associated infections. The increasing prevalence of antimicrobial-resistant strains, particularly methicillin-resistant *S. aureus* (MRSA), has complicated treatment and increased the need for reliable facility-specific antimicrobial resistance surveillance. However, limited published data are available on the prevalence and antimicrobial susceptibility patterns of *S. aureus* at Meru Teaching and Referral Hospital (MeTRH), a major referral facility in eastern Kenya.

**Objective:** To determine the prevalence of *S. aureus* among clinical specimens processed at MeTRH and assess the antimicrobial susceptibility patterns and proportion of MRSA among confirmed isolates.

**Methods:** A retrospective descriptive cross-sectional study was conducted using microbiology laboratory records for clinical specimens submitted for bacteriological culture at MeTRH during the study period. A census approach was used, whereby all eligible laboratory records were included. Clinical specimens were processed using routine microbiological procedures, and suspected *S. aureus* isolates were identified using conventional methods and the VITEK® 2 Compact automated identification system. Antimicrobial susceptibility testing was performed using VITEK® 2 Compact, with interpretation based on Clinical and Laboratory Standards Institute criteria. Methicillin resistance was determined phenotypically using cefoxitin susceptibility as the surrogate marker. Data were analysed using frequencies and percentages.

**Results:** A total of 1,082 clinical specimens were analysed, of which 37 yielded *S. aureus*, giving an overall prevalence of 3.42%. Among the confirmed isolates, 21 (56.8%) were recovered from male patients and 16 (43.2%) from female patients. Pus was the predominant specimen type, accounting for 25 (67.6%) isolates, followed by other specimens at 8 (21.6%) and urine at 4 (10.8%). Antimicrobial susceptibility varied considerably. Resistance was highest to benzylpenicillin and amoxicillin, with 100% resistance among tested isolates. Resistance to amoxicillin-clavulanic acid was 75.0%, while 63.3% of isolates tested were resistant to oxacillin. Susceptibility was relatively high to gentamicin (82.8%), ciprofloxacin (77.3%) and clindamycin (75.0%). All isolates tested against linezolid and vancomycin were susceptible. Among 30 isolates with methicillin susceptibility results, 19 (63.3%) were classified as MRSA and 11 (36.7%) as methicillin-susceptible *S. aureus* (MSSA).

**Conclusion:** The prevalence of *S. aureus* among clinical specimens processed at MeTRH was relatively low; however, a substantial proportion of isolates demonstrated antimicrobial resistance, particularly to commonly used β-lactam antibiotics. The high proportion of MRSA is of clinical and public health importance. Routine antimicrobial susceptibility testing, hospital-specific antibiograms, antimicrobial stewardship and strengthened infection prevention and control measures are recommended to support appropriate treatment and limit the spread of resistant *S. aureus*.

## Introduction

*Staphylococcus aureus* is a Gram-positive, facultatively anaerobic coccus that commonly colonizes the skin and anterior nares of healthy individuals. Although colonization is frequently asymptomatic, *S. aureus* can cause a broad spectrum of infections ranging from superficial skin and soft tissue infections to severe diseases including bacteraemia, pneumonia, osteomyelitis, endocarditis and sepsis. Its ability to acquire antimicrobial resistance has made it an important pathogen in both community and healthcare settings.

Antimicrobial resistance among *S. aureus* has become an important public health concern. Resistance to β-lactam antibiotics, particularly penicillins and methicillin, substantially reduces the number of effective therapeutic options. Methicillin-resistant *S. aureus* is especially important because resistance is generally associated with the acquisition of the *mecA* or related resistance determinants, resulting in resistance to most β-lactam antimicrobial agents. MRSA infections may be associated with prolonged hospitalization, increased treatment costs, treatment failure and increased morbidity.

The epidemiology and antimicrobial susceptibility of *S. aureus* vary considerably between geographical regions, healthcare facilities, patient populations and specimen types. Consequently, facility-specific surveillance is important for developing reliable antibiograms and guiding empirical antimicrobial therapy. The availability of local susceptibility data is particularly important in resource-limited settings where empirical prescribing may occur before definitive laboratory results are available.

In Kenya, antimicrobial resistance surveillance has been identified as a national priority. The Kenya National Action Plan on Prevention and Containment of Antimicrobial Resistance emphasizes strengthening laboratory-based surveillance, antimicrobial stewardship, infection prevention and control, and evidence-informed antimicrobial prescribing.

Meru Teaching and Referral Hospital is a major public referral facility serving Meru County and neighbouring counties in the Upper Eastern region of Kenya. Its microbiology laboratory receives a broad range of clinical specimens and performs routine bacterial culture, organism identification and antimicrobial susceptibility testing. Despite the importance of *S. aureus* as a clinical pathogen, facility-specific information on its prevalence, antimicrobial susceptibility patterns and MRSA burden remains limited.

This study therefore sought to generate local evidence by determining the prevalence of *S. aureus* among clinical specimens processed at MeTRH and describing the antimicrobial susceptibility patterns and proportion of MRSA among confirmed isolates.

## Study Objectives

### General Objective

To determine the prevalence and antimicrobial susceptibility patterns of *Staphylococcus aureus* isolates from clinical samples at MeTRH laboratory.

### Specific Objectives

1. To determine the prevalence of *S. aureus* isolates over the study period at MeTRH laboratory.
2. To assess the antimicrobial susceptibility patterns of these isolates to commonly used antibiotics and the proportion of MRSA among the isolates.

## Materials and Methods

### Study Design and Setting

A hospital-based retrospective descriptive cross-sectional study was conducted at Meru Teaching and Referral Hospital, a Level 5 public referral hospital in Meru County, Kenya. The study was undertaken within the Department of Medical Laboratory Services, specifically the Microbiology Laboratory.

The laboratory routinely processes clinical specimens including blood, urine, pus, wound swabs, high vaginal swabs, sputum, tissue, aspirates and other body fluids. Bacterial identification and antimicrobial susceptibility testing are performed using standardized laboratory procedures. The laboratory operates within a quality management system and participates in internal quality control and external quality assessment programmes.

### Study Population

The study population comprised clinical specimens submitted to the MeTRH Microbiology Laboratory for bacteriological culture during the retrospective study period. The specimens originated from both inpatient and outpatient departments and represented patients of different age groups and both sexes.

All eligible clinical specimens constituted the denominator for determining the prevalence of *S. aureus*. Specimens yielding confirmed *S. aureus* isolates constituted the analytical population for antimicrobial susceptibility analysis.

### Sampling and Eligibility

A census sampling approach was used. All eligible laboratory records available during the retrospective study period were reviewed rather than selecting a sample. This approach maximized the use of available laboratory information and minimized sampling error.

Eligible records included clinical specimens submitted for bacteriological culture with sufficient demographic, specimen and microbiological information and confirmed *S. aureus* isolates identified through routine laboratory procedures. Duplicate isolates from the same patient and specimen type were excluded to minimize overestimation.

### Specimen Processing and Identification

Clinical specimens were processed according to routine microbiology laboratory standard operating procedures. Depending on specimen type, samples were inoculated onto appropriate culture media, including Blood Agar, Chocolate Agar, MacConkey Agar and Cystine-Lactose-Electrolyte-Deficient (CLED) agar.

Suspected bacterial colonies were evaluated using colony morphology and conventional microbiological tests. Suspected *S. aureus* isolates were subjected to Gram staining, catalase testing and coagulase testing. Final identification was performed using the VITEK® 2 Compact automated identification system. Only isolates confirmed as *S. aureus* were included in the antimicrobial susceptibility analysis.

### Antimicrobial Susceptibility Testing

Antimicrobial susceptibility testing was performed using the VITEK® 2 Compact automated system. Results were interpreted according to applicable Clinical and Laboratory Standards Institute (CLSI) criteria.

The antimicrobial agents evaluated included commonly tested β-lactams, aminoglycosides, fluoroquinolones, macrolides, lincosamides, glycopeptides, oxazolidinones and tetracyclines.

Because antimicrobial panels varied according to the isolates and laboratory testing requirements, not all isolates were tested against every antimicrobial agent. Consequently, denominators varied between antimicrobial agents.

### Determination of MRSA

Methicillin resistance was determined phenotypically using susceptibility to cefoxitin as the surrogate marker in accordance with CLSI recommendations. Isolates classified as resistant to the methicillin surrogate were categorized as MRSA, while susceptible isolates were categorized as MSSA. The study did not undertake molecular detection of resistance genes.

## Data Management and Analysis

Data were extracted from microbiology laboratory records using a standardized data abstraction tool. Data were checked for completeness, consistency, duplication and logical errors before analysis.

The prevalence of *S. aureus* was calculated as the proportion of clinical specimens yielding confirmed *S. aureus* isolates:

Prevalence (%) = Number of *S. aureus* isolates / Total clinical specimens processed × 100

Antimicrobial susceptibility results were summarized using frequencies and percentages for susceptible, intermediate and resistant categories. The proportion of MRSA was calculated among isolates for which methicillin susceptibility results were available.

Statistical analysis was performed using SPSS version 29.

## Ethical Considerations

Ethical approval was obtained from the relevant research and institutional review authorities. Patient confidentiality was maintained through the use of unique identification codes and exclusion of personal identifiers from the analytical dataset. Access to laboratory information was restricted to the research team.

## Results

### Prevalence of *Staphylococcus aureus*

A total of 1,082 clinical specimens were analysed during the retrospective study. Of these, 37 specimens yielded *S. aureus*, giving an overall prevalence of 3.42%. The remaining 1,045 specimens (96.58%) either yielded other organisms or had no *S. aureus* isolated.

**Table 1.**
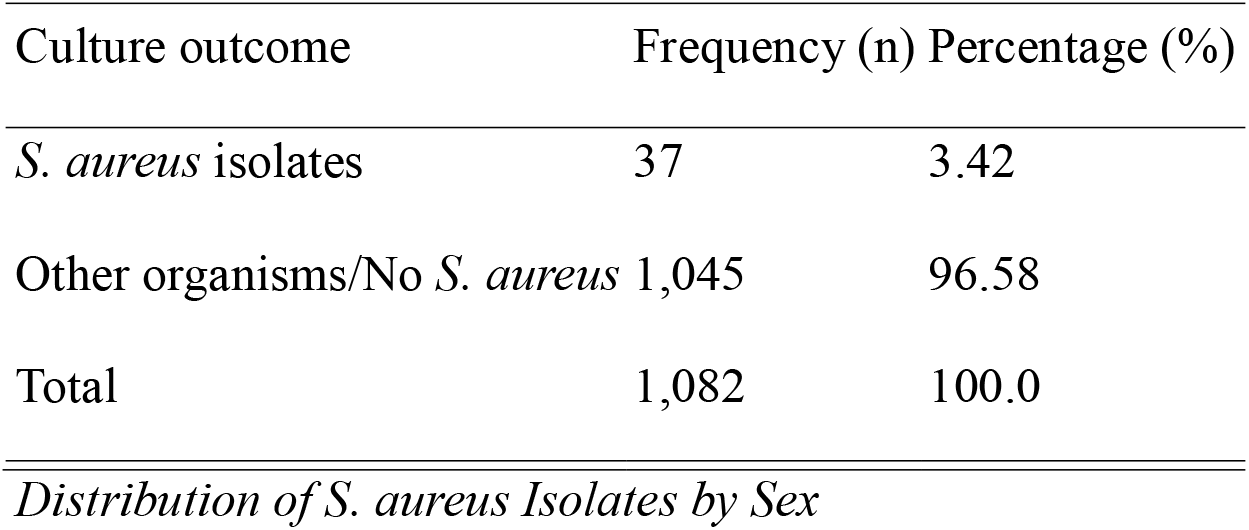
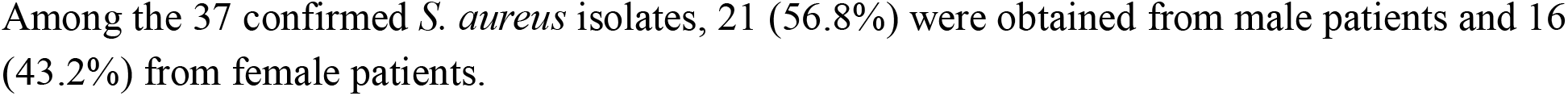
Prevalence of *Staphylococcus aureus* among clinical specimens.

**Table 2.**
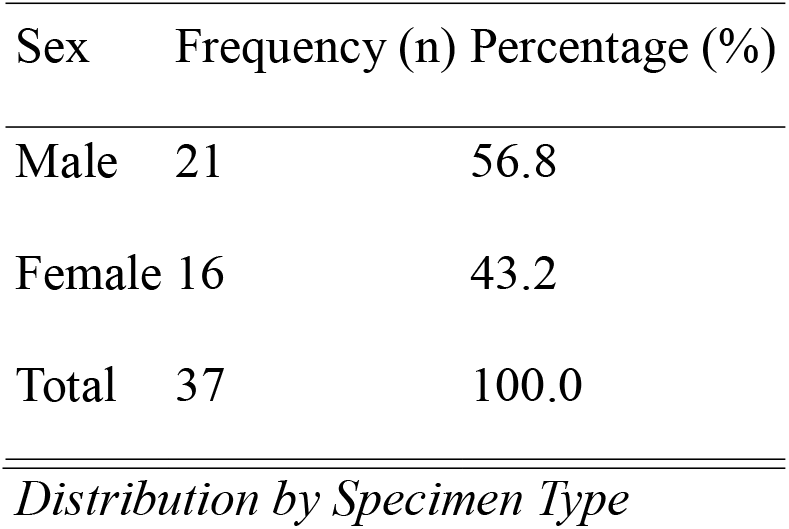
Distribution of *S. aureus* isolates by sex.

**Table 3.** Distribution of *S. aureus* isolates by specimen type.

| Specimen type | Frequency (n) | Percentage (%) |
| --- | --- | --- |
| Pus | 25 | 67.6 |
| Other specimens | 8 | 21.6 |
| Urine | 4 | 10.8 |
| Total | 37 | 100.0 |

**Table 4.** Antimicrobial susceptibility profile of *S. aureus* isolates.

| Antimicrobial agent | Tested (n) | Susceptible n (%) | Intermediate n (%) | Resistant n (%) |
| --- | --- | --- | --- | --- |
| Benzylpenicillin | 18 | 0 (0.0) | 0 (0.0) | 18 (100.0) |
| Amoxicillin | 5 | 0 (0.0) | 0 (0.0) | 5 (100.0) |
| Amoxicillin-clavulanic acid | 4 | 0 (0.0) | 1 (25.0) | 3 (75.0) |
| Oxacillin | 30 | 11 (36.7) | 0 (0.0) | 19 (63.3) |
| Gentamicin | 29 | 24 (82.8) | 0 (0.0) | 5 (17.2) |
| Ciprofloxacin | 22 | 17 (77.3) | 0 (0.0) | 5 (22.7) |
| Clindamycin | 16 | 12 (75.0) | 0 (0.0) | 4 (25.0) |
| Erythromycin | 17 | 8 (47.1) | 0 (0.0) | 9 (52.9) |
| Tetracycline | 16 | 9 (56.3) | 0 (0.0) | 7 (43.7) |
| Linezolid | 13 | 13 (100.0) | 0 (0.0) | 0 (0.0) |
| Vancomycin | 12 | 12 (100.0) | 0 (0.0) | 0 (0.0) |

*Percentages are calculated using the number of isolates tested against each antimicrobial agent.*

### Antimicrobial Susceptibility Patterns

Marked variation in antimicrobial susceptibility was observed among the *S. aureus* isolates.

Complete resistance was observed among isolates tested against benzylpenicillin and amoxicillin. Of the 30 isolates tested against oxacillin, 19 (63.3%) were resistant and 11 (36.7%) were susceptible.

Gentamicin demonstrated the highest susceptibility among the commonly tested agents, with 24 of 29 isolates (82.8%) susceptible. Ciprofloxacin susceptibility was 77.3%, while clindamycin susceptibility was 75.0%.

Resistance to erythromycin was 52.9%, while tetracycline resistance was 43.7%. All isolates tested against linezolid and vancomycin were susceptible.

### Antimicrobial Resistance Ranking

The highest resistance was observed against benzylpenicillin and amoxicillin (100.0% each), followed by amoxicillin-clavulanic acid (75.0%) and oxacillin (63.3%). Erythromycin resistance was 52.9%, while tetracycline resistance was 43.7%.

Resistance was lower for clindamycin (25.0%), ciprofloxacin (22.7%) and gentamicin (17.2%). No resistance was observed among isolates tested against linezolid and vancomycin.

**Table 5.** Ranking of antimicrobial resistance.

| Rank | Antimicrobial agent | Resistance (%) |
| --- | --- | --- |
| 1 | Benzylpenicillin | 100.0 |
| 1 | Amoxicillin | 100.0 |
| 3 | Amoxicillin-clavulanic acid | 75.0 |
| 4 | Oxacillin | 63.3 |
| 5 | Erythromycin | 52.9 |
| 6 | Tetracycline | 43.7 |
| 7 | Clindamycin | 25.0 |
| 8 | Ciprofloxacin | 22.7 |
| 9 | Gentamicin | 17.2 |
| 10 | Linezolid | 0.0 |
| 10 | Vancomycin | 0.0 |

### Prevalence of Methicillin-Resistant *Staphylococcus aureus*

Oxacillin susceptibility results were available for 30 isolates and were used to classify the isolates as MRSA or MSSA.

Of the 30 isolates, 19 (63.3%) were classified as MRSA, while 11 (36.7%) were classified as MSSA. Thus, nearly two-thirds of the isolates with available methicillin susceptibility results demonstrated phenotypic methicillin resistance.

**Table 6.** Distribution of MRSA and MSSA isolates.

| Classification | Frequency (n) | Percentage (%) |
| --- | --- | --- |
| MRSA | 19 | 63.3 |
| MSSA | 11 | 36.7 |
| Total | 30 | 100.0 |

## Discussion

This study found an overall *S. aureus* prevalence of 3.42% among 1,082 clinical specimens processed at MeTRH. The relatively low proportion indicates that *S. aureus* accounted for a small proportion of all clinical specimens submitted for bacteriological investigation. However, the public health importance of the organism should not be judged solely by its prevalence because a substantial proportion of the recovered isolates demonstrated antimicrobial resistance.

The observed prevalence is influenced by the broad range of specimens included in the study. Studies restricted to wound or skin and soft tissue infections may report substantially higher *S. aureus* prevalence because the organism is an important cause of abscesses, wound infections and surgical-site infections. In the present study, all clinical specimens submitted for routine bacteriological investigation were considered, resulting in a broader denominator.

The distribution of isolates showed a modest male predominance, with males accounting for 56.8% of confirmed isolates. This pattern may partly reflect differences in exposure to trauma, occupational activities, surgical procedures or healthcare utilization. However, the study was not designed to establish causal associations between sex and *S. aureus* infection.

Pus was the predominant specimen type, accounting for 67.6% of all *S. aureus* isolates. This finding is consistent with the established role of *S. aureus* as an important cause of skin and soft tissue infections, abscesses and wound infections. The predominance of pus isolates also has implications for infection prevention and clinical management, particularly within surgical and wound-care services.

The antimicrobial susceptibility findings are of considerable clinical importance. Resistance to benzylpenicillin and amoxicillin was 100% among the isolates tested. This finding is consistent with the long-standing predominance of β-lactamase-mediated penicillin resistance among *S. aureus*. The high resistance rate indicates that these agents should not be relied upon for empirical treatment of suspected *S. aureus* infections at the facility without supporting susceptibility results.

Oxacillin resistance was observed in 63.3% of tested isolates. This high level of resistance corresponds to the substantial MRSA burden identified in the study. MRSA is clinically important because resistance to methicillin is generally associated with resistance to most β-lactam antibiotics, thereby substantially narrowing therapeutic choices.

The relatively high susceptibility to gentamicin (82.8%), ciprofloxacin (77.3%) and clindamycin (75.0%) indicates that these agents retained activity against a considerable proportion of isolates. Nevertheless, susceptibility results should be interpreted at the individual-isolate level and used to guide definitive therapy rather than assuming universal effectiveness.

Resistance to erythromycin was relatively high at 52.9%, while tetracycline resistance was 43.7%. These findings demonstrate that commonly used oral antimicrobial agents may have limited effectiveness against a considerable proportion of *S. aureus* isolates. Consequently, empirical treatment should take into account local susceptibility patterns.

All isolates tested against linezolid and vancomycin were susceptible. This finding is encouraging because these agents remain important therapeutic options for serious infections caused by MRSA. However, their continued effectiveness should be protected through antimicrobial stewardship because unnecessary exposure to reserve agents may promote the emergence of resistance.

The MRSA proportion of 63.3% is particularly important for MeTRH. The high burden may reflect several factors, including antimicrobial selection pressure, previous antibiotic exposure, prolonged hospitalization, referral of patients with complicated infections and healthcare-associated transmission. However, these factors were not directly measured in the present study and therefore causal explanations cannot be established.

The findings emphasize the importance of facility-specific antimicrobial surveillance. National or regional resistance estimates may not accurately reflect the susceptibility profile of individual hospitals. Regular hospital antibiograms can provide clinicians with locally relevant information for empirical therapy while supporting antimicrobial stewardship and infection prevention and control activities.

### Clinical and Public Health Implications

The high resistance to commonly used β-lactam antibiotics has important implications for empirical management of suspected *S. aureus* infections at MeTRH. Culture and antimicrobial susceptibility testing should be prioritized whenever clinically feasible, particularly in patients with severe, recurrent or healthcare-associated infections.

The substantial MRSA burden also emphasizes the need for strengthened infection prevention and control measures. These should include consistent hand hygiene, appropriate environmental cleaning, adherence to standard precautions, appropriate management of infected wounds and timely identification of patients with suspected or confirmed resistant infections.

The findings also support the development and periodic updating of a hospital-specific *S. aureus* antibiogram. Such an antibiogram can assist clinicians and antimicrobial stewardship teams in selecting appropriate empirical therapy while reducing unnecessary exposure to ineffective antibiotics.

### Limitations

This study has several limitations. First, it was conducted at a single referral hospital, which may limit the generalizability of the findings to other healthcare facilities or the wider community.

Second, the retrospective nature of the study meant that the analysis depended on the completeness and accuracy of routinely collected laboratory records. Although data quality checks were undertaken, some clinical information was unavailable.

Third, not all isolates were tested against every antimicrobial agent, resulting in different denominators for individual antibiotics. Therefore, susceptibility percentages should be interpreted in relation to the number of isolates tested for each antimicrobial.

Fourth, the study did not assess important patient-related factors such as previous antibiotic exposure, comorbidities, duration of hospitalization or clinical outcomes. Consequently, factors associated with antimicrobial resistance could not be determined.

Finally, MRSA was identified using phenotypic susceptibility testing rather than molecular detection of resistance genes. The findings therefore describe phenotypic methicillin resistance and do not provide information on the underlying molecular mechanisms.

## Conclusion

The retrospective review demonstrated an overall *Staphylococcus aureus* prevalence of 3.42% among 1,082 clinical specimens processed at Meru Teaching and Referral Hospital. Pus was the predominant source of isolates, accounting for 67.6%, while males accounted for 56.8% of confirmed isolates.

The antimicrobial susceptibility profile demonstrated substantial resistance to commonly used β-lactam antibiotics, with 100% resistance to benzylpenicillin and amoxicillin among tested isolates and 63.3% resistance to oxacillin. In contrast, gentamicin, ciprofloxacin and clindamycin retained relatively good activity, while all tested isolates remained susceptible to linezolid and vancomycin.

A substantial MRSA burden was identified, with 63.3% of isolates with available methicillin susceptibility results classified as MRSA. This finding highlights the importance of continuous antimicrobial resistance surveillance, routine culture and susceptibility testing, antimicrobial stewardship and strengthened infection prevention and control measures at MeTRH.

### Recommendations

1. Routine antimicrobial susceptibility testing should continue for clinically significant *S. aureus* isolates to support targeted antimicrobial therapy.
2. Hospital-specific antibiograms should be generated and periodically updated to guide empirical antimicrobial prescribing.
3. Antibiotics demonstrating very high resistance, particularly benzylpenicillin, amoxicillin and oxacillin, should not be routinely relied upon for empirical treatment of suspected *S. aureus* infections without microbiological evidence.
4. Antimicrobial stewardship interventions should promote culture-guided prescribing and prudent use of reserve agents such as vancomycin and linezolid.
5. Infection prevention and control measures should be strengthened, particularly hand hygiene, environmental cleaning and appropriate management of patients with suspected or confirmed MRSA.
6. Further studies should investigate the molecular epidemiology of MRSA and associated resistance determinants to complement phenotypic surveillance.

## Data Availability

all data produced in the present study are available upon reasonable request to the authors.

https://www.pharis.com

## Declarations

### Ethics Approval and Consent to Participate

Ethical approval was obtained from the relevant institutional and county research authorities before commencement of the study. The study used routinely collected laboratory records, and patient identifiers were excluded from the analytical dataset.

### Consent for Publication

Not applicable.

### Availability of Data

The data supporting the findings are derived from laboratory records at Meru Teaching and Referral Hospital. Access to patient-level information is subject to institutional confidentiality and ethical requirements.

### Author Contributions

Pharis Kaumbuthu Mutia conceived the study, contributed to data collection and laboratory data interpretation, performed the analysis, and prepared the manuscript. Dorothy Kagendo and Jane Rutto, contributed to the study supervision, laboratory/research activities, interpretation of findings, and critical review of the manuscript.

## References

1. Baur D, Gladstone BP, Burkert F, et al. Effect of antibiotic stewardship on the incidence of antimicrobial resistance and Clostridium difficile infection: a systematic review and meta-analysis. The Lancet Infectious Diseases. 2017;17:990–1001.

2. Cheung GYC, Bae JS, Otto M. Pathogenicity and virulence of Staphylococcus aureus. Virulence. 2021;12(1):547–569.

3. Clinical and Laboratory Standards Institute. Performance Standards for Antimicrobial Susceptibility Testing. 35th ed. CLSI supplement M100. Wayne, PA: Clinical and Laboratory Standards Institute; 2025.

4. Gitau W, Masika MM, Waiyaki PG, et al. Antimicrobial susceptibility patterns among Staphylococcus aureus isolates from clinical specimens in Kenya.

5. Lakhundi S, Zhang K. Methicillin-resistant Staphylococcus aureus: molecular characterization, evolution, and epidemiology. Clinical Microbiology Reviews. 2018;31(4):e00020–18.

6. Laxminarayan R, Van Boeckel T, Frost I, et al. The Lancet Infectious Diseases Commission on antimicrobial resistance. The Lancet Infectious Diseases. 2020.

7. Lee AS, de Lencastre H, Garau J, et al. Methicillin-resistant Staphylococcus aureus. Nature Reviews Disease Primers. 2018;4:18033.

8. Ministry of Health, Kenya. National Action Plan for Prevention and Containment of Antimicrobial Resistance 2023–2027. Nairobi: Ministry of Health; 2023.

9. Murray CJL, Ikuta KS, Sharara F, et al. Global burden of bacterial antimicrobial resistance in 2019: a systematic analysis. The Lancet. 2022;399:629–655.

10. World Health Organization. Global antimicrobial resistance and use surveillance system (GLASS) report. Geneva: World Health Organization; 2024.

